# Epidemiological and Clinical Features, Treatment Status, and Economic Burden of Traumatic Spinal Cord Injury in China

**DOI:** 10.1101/2021.09.27.21264179

**Authors:** Hengxing Zhou, Yongfu Lou, Lingxiao Chen, Yi Kang, Lu Liu, Zhiwei Cai, David B Anderson, Wei Wang, Chi Zhang, Jinghua Wang, Guangzhi Ning, Yanzheng Gao, Baorong He, Wenyuan Ding, Yisheng Wang, Wei Mei, Yueming Song, Yue Zhou, Maosheng Xia, Huan Wang, Jie Zhao, Guoyong Yin, Tao Zhang, Feng Jing, Rusen Zhu, Bin Meng, Li Duan, Zhongliang Deng, Zhongmin Zhang, Desheng Wu, Yajun Liu, Zhengdong Cai, Lin Huang, Zhanhai Yin, Kainan Li, Shibao Lu, Xu Cao, Shiqing Feng

## Abstract

**Background:** China has the largest population of traumatic spinal cord injury (TSCI), but has not yet performed a national-level study on its epidemiological and clinical features, treatment status, and economic burden.

**Methods:** A total of 14 754 patients were recruited between January 2013 and December 2018 from 37 hospitals in 11 provinces and municipalities, which represented all geographical divisions of China. The percentage of TSCI in hospitalized patients and the percentage of TSCI in hospitalized patients through the orthopaedic departments were calculated. The treatment status, total and daily costs were collected.

**Results:** The percentage of TSCI in hospitalized patients and the percentage of TSCI in hospitalized patients through the orthopaedic departments did not change significantly overall (APC= -0.5%, 95% CI: -3.0 to 2.1 and -1.6%, -4.9 to 1.8, respectively). A total of 10 918 (74.0%) patients received surgery after TSCI. However, only 3.0% of patients underwent surgery received surgery less than 24 hours after injury. A total of 2 084 (14.1%) patients were treated with methylprednisolone sodium succinate/methylprednisolone (MPSS/MP) at a high dose (≥500 mg) and 641 (4.3%) patients receiving it within 8 hours. The total costs for acute TSCI decreased (-4.8%, -6.2 to -3.4), while the daily costs did not change significantly (0.5%, -1.2 to 2.2).

**Conclusions:** This study revealed epidemiological and clinical features, treatment status, and economic burden of TSCI that occurred in China from 2013 to 2018.

**Funding:** National Key Research and Development Project of Stem Cell and Transformation Research (2019YFA0112100).

## Introduction

Traumatic spinal cord injury (TSCI) is a significant life event which can lead to permanent disability or even death *(Ahuja et al., 2017; National Spinal Cord Injury Statistical Center, 2016)*. TSCI can be treated with surgery, glucocorticoids or other non-surgery treatments; followed by a period of rehabilitation *(Robert R Hansebout, 2021)*. These treatments bring a heavy financial burden to patients and their families, with direct costs ranging from US$ 1.1 to 4.7 million per patient over their lifetime *(National Spinal Cord Injury Statistical Center, 2016)*. Over the past three decades, there has been an increased understanding of the burden of TSCI globally, attributable to epidemiological studies, such as the Global Burden of Disease (GBD) Study *(GBD 2016 Traumatic Brain Injury and Spinal Cord Injury Collaborators, 2019)*. The GBD studies concluded that the number of people with TSCI would increase globally *(Institute for Health Metrics and Evaluation, 2021)*. The GBD study did report limitations with the accuracy of data in some regions however, and did not collect data on the severity of TSCI, limiting calculations on disease burden *(Badhiwala et al., 2019)*. Authors of the GBD study recommended more researches be conducted into developing countries and the collection of more comprehensive TSCI data *(Badhiwala et al., 2019)*. Currently, the high-quality research in developing countries is still limited, especially in China which is one developing country of significance, given it possesses the largest population of any country and therefore has capacity to significantly influence global rates of TSCI *(Institute for Health Metrics and Evaluation, 2021)*. One major limitation of the current data available from China is that most of them are from regional areas *(Yuan et al., 2018)*.

The purpose of this study was to identify the epidemiological and clinical features, treatment status, and economic burden of TSCI in China from 2013 to 2018.

## Methods

We followed STrengthening the Reporting of OBservational studies in Epidemiology (STROBE) statement *(von Elm et al., 2007)*.

### Study population

We established the Chinese multicentre TSCI registry project (CMTSCIRP, ChiCTR1800019691) which was a national, hospital-based retrospective study. Any patients who sustained a TSCI between January 2013 and December 2018 in China and attended a large general hospital or orthopaedic speciality centre were included. The sites included 37 hospitals from the seven geographical regions (Central China, Northern China, Eastern China, Southern China, Northwest China, Southwest China, and Northeast China) and four municipalities (Beijing, Shanghai, Tianjin, and Chongqing) of China. For each region or municipality, we selected between three and four hospitals that were either large general hospitals or orthopaedic speciality centres (Figure supplement 1). The study was approved by Medical Research Ethics Committee of Tianjin Medical University General Hospital [IRB2018-154-01] and all participating centres.

Spinal cord injury (SCI) is defined as damage to the spinal cord that temporarily or permanently causes its functional changes, and TSCI occurs when an external physical impact (such as falls, motor vehicle injuries, or sports-related injuries) acutely damages the spinal cord *(Ahuja et al., 2017)*.

With the help of the librarian, we used the following strategy to retrieve TSCI cases. We identified TSCI mainly using medical terms in Chinese ("Spinal fractures", "Cervical fractures", "Thoracic fractures", "Lumbar fractures", "Sacral fractures", "Spinal cord injury", "Cervical spine fracture with spinal cord injury", "Thoracic spine fracture with spinal cord injury", "Lumbar spine fracture with spinal cord injury", "Cervical spinal cord injury", "Thoracic spinal cord injury", "Lumbar spinal cord injury", "Cauda equina injury", "Nerve injury", "Quadriplegia", "Paraplegia", "Incomplete quadriplegia", and "Incomplete paraplegia"), as it was difficult to unify case codes due to diversities in codes used by different hospitals. And the Tenth Revision of the International Classification of Diseases (ICD-10) codes involved in TSCI from the International Spinal Cord Injury Community Survey (InSCI) Group was also used to complement and refine the medical terms [S12 (Fracture of the neck), S12.0 (Fracture of first cervical vertebra), S12.2 (Fracture of other specified cervical vertebra), S13.0 (Traumatic rupture of cervical intervertebral disk), S13.2 (Dislocation of other and unspecified parts of neck), S13.4 (Sprain and strain of cervical spine), S14 (Injury of nerves and spinal cord at neck level), S14.0 (Concussion and edema of cervical spinal cord), S14.1 (Other and unspecified injuries of cervical spinal cord), S17 (Crushing injury of neck), S19 (Other and unspecified injuries of neck), S22 (Fracture of ribs, sternum and thoracic spine), S22.0 (Fracture of thoracic vertebra), S23.1 (Dislocation of thoracic vertebra), S24 (Injury of nerves and spinal cord at thorax level), S24.0 (Concussion and edema of thoracic spinal cord), S24.1 (Other and unspecified injuries of thoracic spinal cord), S28 (Crushing injury of thorax and traumatic amputation of part of thorax), S29 (Other and unspecified injuries of thorax), S32 (Fracture of lumbar spine and pelvis), S32.0 (Fracture of lumbar vertebra), S33.1 (Dislocation of lumbar vertebra), S34.0 (Concussion and edema of lumbar spinal cord), S34.1 (Other injury of lumbar spinal cord), S34.3 (Injury of cauda equina), S38 (Crushing injury and traumatic amputation of part of abdomen, lower back and pelvis), S39 (Other and unspecified injuries of abdomen,lower back and pelvis), T02.0 (Fractures involving head with neck), T02.1 (Fractures involving thorax with lower back and pelvis), T04.1 (Crushing injuries involving thorax with abdomen,lower back and pelvis), T04.2 (Crushing injuries involving multiple regions of upper limbs), T06.0 (Injuries of brain and cranial nerves with injuries of nerves and spinal cord at neck level), T06.1 (Injuries of nerves and spinal cord involving other multiple body regions), T09.3 (Injury of spinal cord, level unspecified), T09.4 (Injury of unspecified nerve, spinal nerve root and plexus of trunk), T91.1 (Sequelae of fracture of spine), T91.3 (Sequelae of injury of spinal cord), G82 (Paraplegia and tetraplegia), G82.0 (Flaccid paraplegia), G82.1 (Spastic paraplegia), G82.2 (Paraplegia, unspecified), G82.3 (Flaccid tetraplegia), G82.4 (Spastic tetraplegia), G82.5 (Tetraplegia, unspecified), G83.4 (Cauda equina syndrome)] *(Gross-Hemmi et al., 2017)*. All cases were checked by trained investigators. Non-traumatic spinal cord injury (such as tumor or degenerative compression) was excluded.

### Outcomes

Considering the accessibility of data, the percentage of TSCI in hospitalized patients of 23 hospitals were calculated from: the number of TSCI patients/the number of hospital admissions. We also calculated the percentage of TSCI in hospitalized patients through the orthopaedic departments from the same 23 hospitals as: the number of TSCI patients/the number of orthopaedic departments admissions.

According to the definition for early surgery (<24 h) mentioned in previous studies *(Robert R Hansebout, 2021; Badhiwala et al., 2021)* and the interquartile distribution of data in this study, surgery timing was divided to two sections: 1. "<24 h"; 2. "<4.0 d", "4.0-11.9 d" followed by ">11.9 d". Off-label use of methylprednisolone sodium succinate/methylprednisolone (MPSS/MP) for TSCI remains contentious *(Roberts et al., 2017)*, therefore, we defined 500mg of MPSS/MP as a threshold in this study, based on the user guide *(DXY Drugs Information, 2021)*. The use of MPSS/MP was divided into continuous and intermittent use. Intermittent use referred to that two or more prescriptions, with at least one day interval between arbitrary two prescriptions, were made for MPSS / MP use during hospitalization. The length of accumulated time was the sum of all days to use MPSS/MP. Although the use of MPSS/MP after SCI continues to be controversial, AOSpine organization suggested a high-dose of MPSS/MP within 8 hours of acute SCI as a treatment option in 2017 *(Fehlings et al., 2017)*. The starting time to use off-label MPSS/MP (≥500 mg) was divided into "≤8 h", and ">8 h". There rarely has been studies on whether normal dose of MPSS/MP and the starting time affect the treatment for SCI. Therefore, based on the interquartile distribution of data from patients of continuously using MPSS/MP with a normal dose (<500 mg), the starting time of MPSS/MP use was divided into "≤1.6 d", "1.6-9.0 d", and "≥9.0 d". And the length of accumulated use time was divided into "≤2 d", "3-5d", and "≥6 d". In addition, we removed participants (1 case excluded in the high-dose group and 7 cases excluded in the normal dose group) who lacked starting time from injury to medication.

The total and daily costs were the costs during the patient’s hospitalization for acute TSCI and were identified from the patient’s medical record. And acute TSCI was defined as hospital admission within 14 days after spinal cord injury. The total and daily costs were converted to 2013 Chinese Yuan (CNY), discounting at an annual rate of 5%, as recommended by China guidelines for pharmacoeconomic evaluations (2020) *(Gordon, 2020)*. In addition, referring to the exchange rate of CNY to USD (USA dollar, USD=100) in 2013 as CNY 619.32, the costs were showed by CNY and USD *(National Bureau of Statistics, 2021)*. Non-acute TSCI patients were not included in the financial burden of this study, as acute and non-acute TSCI patients were treated differently during hospitalization, which would bias the estimate *(Robert R Hansebout, 2021)*.

### Covariates

Age groups in this study were divided into "15 years∼", "25 years∼","35 years∼", "45 years∼", "55 years∼" and "≥65 years". Occupations covered "Farmers", "Workers", "Retired", "Office clerks", "Students", "Self-employed", "Civil servants", "Drivers", "Teachers" and "Others" ("Unemployed" or "Freelances") based on medical records. The etiological groups consisted of "Low falls" (<1 m), "High falls" (≥1 m), "Traffic accidents", "Struck by falling objects", "Sport related injuries", "Injuries caused by others", "Work related injuries", "Sharp injuries", "Massage related injuries" and "Others" ("Electric shock injuries", "Gunshot wounds", "Crush injuries", "Iatrogenic injuries", etc.). Level of injury comprised "Cervical", "Thoracic", "Lumbosacral", "Multi-site" and "Cauda equina", and "Multi-site" referred to at least two different segments of injury *(Selvarajah et al., 2014)*. Severity was divided into four types: "Complete quadriplegia", "Complete paraplegia", "Incomplete quadriplegia" and "Incomplete paraplegia" based on degree and extent of neurological damage. The American Spinal Injury Association (ASIA) Impairment Scale (AIS), which superseded the original Frankel Scale, has been widely accepted among clinicians and researchers for classification of the location, severity and extent of spinal cord injury, introducing several changes aiming at a more consistent and objective description on the basis of the scores of a standardized examination of myotomes and dermatomes *(Kirshblum et al., 2011)*. Neurological function of TSCI patients admitted was evaluated using the AIS, with a division into complete (AIS grade A) and 4 incomplete (AIS grade B-E) injury grades, and the grades and key description were shown in Table supplement 1 *(Roberts et al., 2017)*. In groups with less than 1% of the total number of patients, they were categorized as "Others" to increase the reproducibility of the results *(Harrell Jr, 2015)*.

### Statistical Analysis

Continuous variables were presented as a mean (standard deviation, SD) and categorical variables as numbers (percentages). Annual percentage change (APC) is usually used to estimate the event change rate during specific calendar years. Joinpoint regression analysis fits a series of joined linear models of the natural logarithm of annual incidence using calendar year as an independent variable, which is often useful to calculate the APC and corresponding 95% confidence interval (95% CI) *(Salomaa et al., 1992)*. Trends in the percentage of TSCI in hospitalized patients, the percentage of TSCI in hospitalized patients through the orthopaedic departments, and costs were assessed by APC, using the following regression model: Log (rt) = a + bt, where Log denoted the natural logarithm and t was the calendar year *(Salomaa et al., 1992)*. The trend (b) was estimated using ordinary regression and 100 × b represented the estimated APC *(Salomaa et al., 1992)*.

As exploratory analyses, we calculated trends of the percentages for each included hospital; we also performed two subgroup analyses based on hospital type (“General hospitals” versus "Orthopaedic hospitals") and economic level ("Above the PCDI [Per Capita Disposable Income]" versus "Below the PCDI") *(National Bureau of Statistics, 2021)*; considering the number of TSCI patients per year admitted by different hospitals varied a lot, we explored the relationship between the number of TSCI per year and the trend of percentages. "Economic level" was divided to "Above the PCDI" (the sum of final consumption expenditure and savings that residents can use, that is, the income that residents can use for free disposal) and "Below the PCDI" according to whether the residents’ PCDI in the municipalities or non-municipality cities the hospitals located was higher than the national PCDI. The national PCDI in 2013 (CNY 18 311) was taken as the reference level. All residents’ PCDIs come from the National Bureau of Statistics of China *(National Bureau of Statistics, 2021)*. We hypothesized that the hospitals’ TSCI number might influence APC *(Joynt et al., 2011)*. Thus, we performed an additional analysis to explore the relationship between the hospitals’ TSCI number and APC. The Joinpoint Regression Program, Version 4.8.0.1-April 2020 (Statistical Methodology and Applications Branch, Surveillance Research Program, National Cancer Institute), was used in the calculation of APC *(National Cancer Institute, 2020)*. Statistical significance was defined as two-tailed P< 0.05. Statistical description of demographic and clinical features, treatment status, and economic burden of TSCI patients was performed using IBM SPSS Statistics Grad Pack 27.0 (IBM SPSS, Chicago, IL, USA). Linear regression and figures drawing were performed using GraphPad Prism version 9.0.2 for Windows (GraphPad Software, San Diego, California USA, www.graphpad.com).

## Results

### Epidemiological and Clinical Features

A total of 14 754 TSCI cases were included in this study, with men accounting for 75.3% (n=11 115). The average age was 50.0 years overall, 49.2 years in men and 52.2 years in women. Considering TSCI by occupations, TSCI patients were more likely to be farmers (5 568, 37.7% of 14 754). The top three etiological causes of TSCI, accounting for 83.9% of all causes were: low falls (4 452, 30.2% of 14 754), high falls (4 398, 29.8% of 14 754), and traffic accidents (3 524, 23.9% of 14 754). TSCI occurred most commonly in the cervical segment (8 944, 60.6% of 14 754) and presented most often as incomplete quadriplegia (6 483, 43.9% of 14 754). Almost half of all patients (7 168, 48.6% of 14 754) were classified as AIS grade D on admission, followed by AIS grade A (3 671, 24.9% of 14 754) and AIS grade C (2 512, 17.0% of 14 754). (Table 1)

**Table 1:** The epidemiological and clinical features among TSCI patients from 2013 to 2018.

| Category | Men | Women | Total |
| --- | --- | --- | --- |
| All cases, n (%) | 11 115 (75.3) | 3 639 (24.7) | 14 754 (100.0) |
| Age, years, mean (SD) | 49.2 (14.3) | 52.2 (15.3) | 50.0 (14.6) |
| Age groups, n (%) |  |  |  |
| 15 years~ | 592 (5.3) | 176 (4.8) | 768 (5.2) |
| 25 years~ | 1 284 (11.6) | 323 (8.9) | 1 607 (10.9) |
| 35 years~ | 2 017 (18.1) | 549 (15.1) | 2 566 (17.4) |
| 45 years~ | 3 139 (28.2) | 960 (26.4) | 4 099 (27.8) |
| 55 years~ | 2 510 (22.6) | 814 (22.4) | 3 324 (22.5) |
| ≥ 65 years | 1 573 (14.2) | 817 (22.5) | 2 390 (16.2) |
| Occupations, n (%) |  |  |  |
| Farmers | 4 244 (38.2) | 1 324 (36.4) | 5 568 (37.7) |
| Workers | 1 504 (13.5) | 271 (7.4) | 1 775 (12.0) |
| Retired | 656 (5.9) | 426 (11.7) | 1 082 (7.3) |
| Office clerks | 530 (4.8) | 203 (5.6) | 733 (5.0) |
| Students | 210 (1.9) | 116 (3.2) | 326 (2.2) |
| Self-employed | 226 (2.0) | 73 (2.0) | 299 (2.0) |
| Civil servants | 147 (1.3) | 70 (1.9) | 217 (1.5) |
| Drivers | 128 (1.2) | 28 (0.8) | 156 (1.1) |
| Others<sup>a</sup> | 2 845 (25.6) | 931 (25.6) | 3 776 (25.6) |
| Missing | 625 (5.6) | 197 (5.4) | 822 (5.6) |
| Etiological groups, n (%) |  |  |  |
| Low falls | 3 109 (28.0) | 1 343 (36.9) | 4 452 (30.2) |
| High falls | 3 500 (31.5) | 898 (24.7) | 4 398 (29.8) |
| Traffic accidents | 2 650 (23.8) | 874 (24.0) | 3 524 (23.9) |
| Struck by falling objects | 763 (6.9) | 118 (3.2) | 881 (6.0) |
| Sport related injuries | 105 (0.9) | 61 (1.7) | 166 (1.1) |
| Others<sup>b</sup> | 641 (5.8) | 243 (6.7) | 884 (6.0) |
| Missing | 347 (3.1) | 102 (2.8) | 449 (3.0) |
| Level of injury, n (%) |  |  |  |
| Cervical | 7 109 (64.0) | 1 835 (50.4) | 8 944 (60.6) |
| Thoracic | 1 695 (15.2) | 678 (18.6) | 2 373 (16.1) |
| Lumbosacral | 1 981 (17.8) | 976 (26.8) | 2 957 (20.0) |
| Multi-site | 125 (1.1) | 58 (1.6) | 188 (1.2) |
| Cauda equina | 21 (0.2) | 8 (0.2) | 29 (0.2) |
| Missing | 184 (1.7) | 84 (2.3) | 268 (1.8) |
| Severity, n (%) |  |  |  |
| Complete quadriplegia | 1 502 (13.5) | 298 (8.2) | 1 800 (12.2) |
| Complete paraplegia | 2 248 (20.2) | 671 (18.4) | 2 919 (19.8) |
| Incomplete quadriplegia | 5 047 (45.4) | 1 436 (39.5) | 6 483 (43.9) |
| Incomplete paraplegia | 2 131 (19.2) | 1 144 (31.4) | 3 275 (22.2) |
| Missing | 187 (1.7) | 90 (2.5) | 277 (1.9) |

| AIS grade<sup>c</sup>, n (%) |  |  |  |
| --- | --- | --- | --- |
| A | 2 933 (26.4) | 738 (20.3) | 3 671 (24.9) |
| B | 803 (7.2) | 206 (5.7) | 1 009 (6.8) |
| C | 1 987 (17.9) | 525 (14.4) | 2 512 (17.0) |
| D | 5 131 (46.2) | 2 037 (56.0) | 7 168 (48.6) |
| E | 151 (1.4) | 97 (2.7) | 248 (1.7) |
| Missing | 110 (1.0) | 36 (1.0) | 146 (1.0) |

<sup>a</sup> "Others" included "Teachers", "Unemployed" and "Freelances". <sup>b</sup> "Others" included "Injuries caused by others", "Work related injuries", "Sharp injuries", "Massage related injuries", "Electric shock injuries", "Gunshot wounds", "Crush injuries", and "Iatrogenic injuries". <sup>c</sup> AIS grade A represents complete impairment with no motor or sensory function below the level of injury; grades B–D represent progressively less severe impairment; AIS grade E represents normal motor and sensory examinations, but still may have abnormal reflexes or other neurologic phenomena. TSCI=traumatic spinal cord injury. AIS=American Spinal Injury Association (ASIA) Impairment Scale. SD=standard deviation.

From 2013 to 2018, the percentage of TSCI in hospitalized patients and the percentage of TSCI in hospitalized patients through the orthopaedic departments did not change significantly overall (APC= -0.5%, 95% CI: -3.0 to 2.1 and APC= -1.6%, 95% CI: -4.9 to 1.8), but the trends varied among individual hospitals (Figure 1 and Figure supplement 2). The percentage of TSCI in hospitalized patients and the percentage of TSCI in hospitalized patients through the orthopaedic departments from general hospitals decreased (APC= -4.2%, 95% CI: -7.2 to -1.0; APC= -5.1%, 95% CI: -8.4 to -1.7) (Table supplement 2). However, the percentage of TSCI in both hospitalized patients (APC= 8.4%, 95% CI: 3.5 to 13.6) and hospitalized patients through the orthopaedic departments (APC= 7 .5%, 95% CI: 2.5 to 12.8) increased when the number of TSCI patients admitted to the hospital per year was greater than 140 (Figure supplement 3).

**Figure 1:**
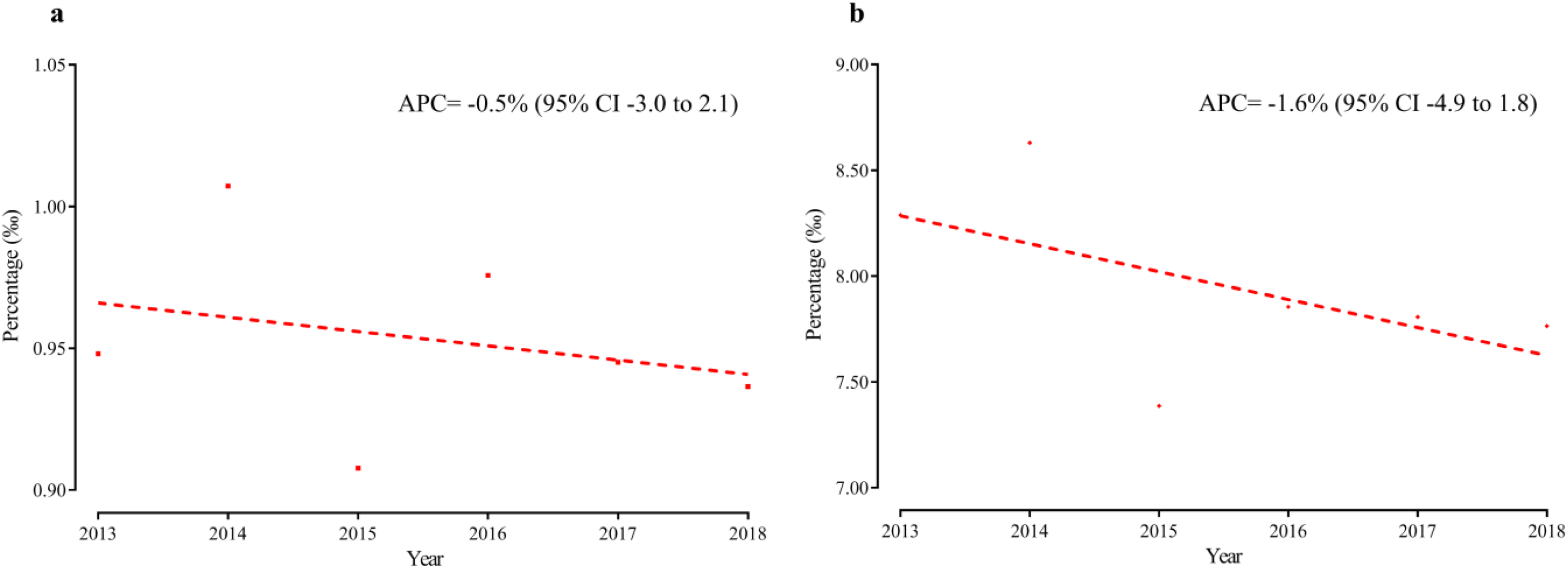
Trends in the percentage among TSCI patients from 2013 to 2018. (a) Trends in the percentage of TSCI in hospitalized patients from 23 hospitals, P= 0.61. (b) Trends in the percentage of TSCI in hospitalized patients through the orthopaedic departments from 23 hospitals, P= 0.25. TSCI=traumatic spinal cord injury. APC=annual percentage change. 95% CI=95% confidence interval.

**Figure 2:**
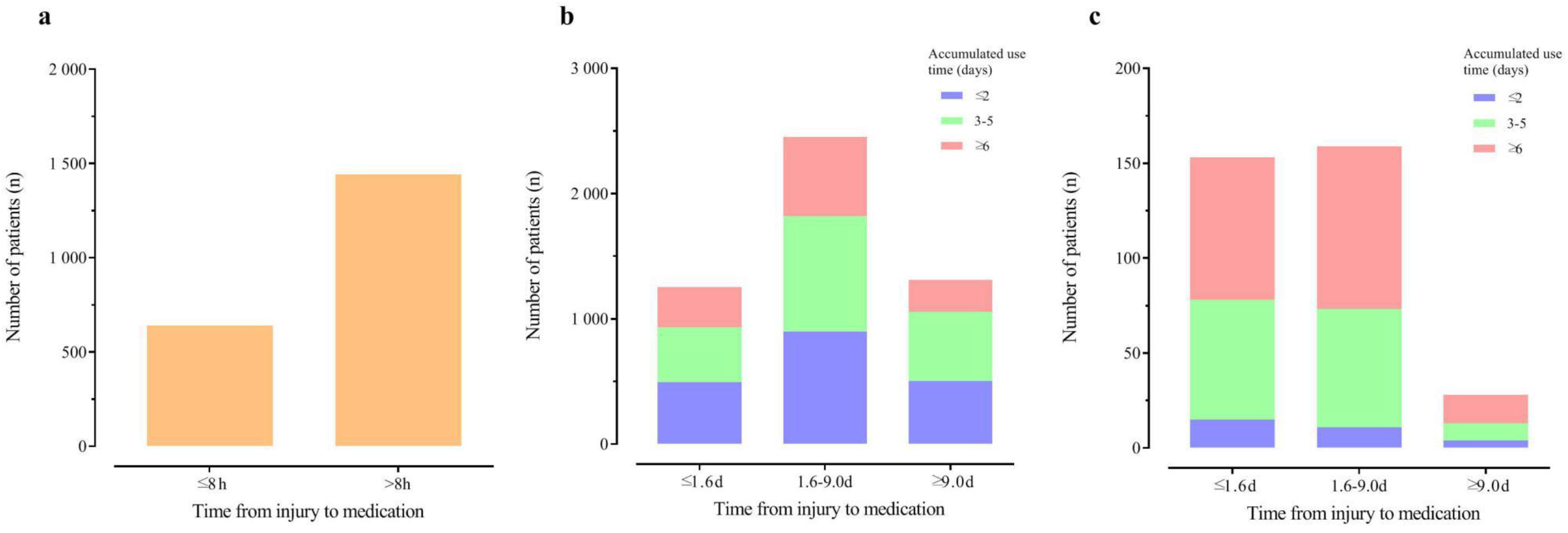
The use of MPSS/MP among TSCI patients during hospitalization. (a) Use of MPSS/MP at over-instruction dose (≥500 mg). (b) Continuous use of MPSS/MP with normal instruction dose (<500 mg). (c) Intermittent use of MPSS/MP with normal instruction dose (<500 mg). MPSS/MP=methylprednisolone sodium succinate/methylprednisolone. TSCI=traumatic spinal cord injury.

## Treatment Status

### Surgery

10 918 (74.0% of 14 754) patients received surgery after TSCI. The highest surgery rate was found in patients aged 15-24 years (591, 77.0% of 768). For etiological groups, patients with high falls were more likely to receive surgery (3 571, 81.2% of 4 398). Considering the injury level, the highest surgery rate was found in patients with cauda equina (27, 93.1% of 29). Patients with complete paraplegia (2 343, 80.3% of 2 919) and AIS grade B (825, 81.8% of 1 009), had the highest surgery rate considering injury severity and AIS grade, respectively. Among the 10 882 patients who had exact surgery timing, a few (324, 3.0% of 10 882) patients underwent surgery in less than 24 hours. In addition, about a quarter (2 698, 24.8% of 10 882) of patients underwent surgery less than 4.0 days, followed by about a half (5 462, 49.9% of 10 882) within 4.0-11.9 days, and a quarter (2 758, 25.3% of 10 882) more than 11.9 days after injury. (Table 2)

**Table 2:** The treatment methods and surgery timing among TSCI patients.

| Category | Treatment methods |  | Surgery timing<sup>a</sup> |  |  |  |
| --- | --- | --- | --- | --- | --- | --- |
|  | Surgery | Conservative therapies | Section 1 | Section 2 |  |  |
|  |  |  | <24 h | <4.0 d | 4.0-11.9 d | >11.9 d |
| Gender, n (%) |  |  |  |  |  |  |
| Men | 8 287 (74.6) | 2 828 (25.4) | 243 (2.9) | 2 038 (24.7) | 4 196 (50.8) | 2 032 (24.6) |
| Women | 2 631 (72.3) | 1 008 (27.7) | 81 (3.1) | 660 (25.2) | 1 230 (47.0) | 726 (27.8) |
| Total | 10 918 (74.0) | 3 836 (26.0) | 324 (3.0) | 2 698 (24.8) | 5 426 (49.9) | 2 758 (25.3) |
| Age groups, n (%) |  |  |  |  |  |  |
| 15 years~ | 591 (77.0) | 177 (23.0) | 26 (4.4) | 177 (30.0) | 301 (51.0) | 112 (19.0) |
| 25 years~ | 1 214 (75.5) | 393 (24.5) | 53 (4.4) | 375 (30.9) | 579 (47.8) | 258 (21.3) |
| 35 years~ | 1 958 (76.3) | 608 (23.7) | 57 (2.9) | 504 (25.9) | 981 (50.4) | 463 (23.8) |
| 45 years~ | 3 098 (75.6) | 1 001 (24.4) | 94 (3.0) | 765 (24.8) | 1 570 (50.8) | 754 (24.4) |
| 55 years~ | 2 474 (74.4) | 850 (25.6) | 67 (2.7) | 570 (23.1) | 1 225 (49.7) | 670 (27.2) |
| ≥ 65 years | 1 583 (66.2) | 807 (33.8) | 27 (1.7) | 307 (19.5) | 770 (48.8) | 501 (31.7) |
| Occupations, n (%) |  |  |  |  |  |  |
| Farmers | 4 212 (75.6) | 1 356 (24.4) | 141 (3.4) | 1 138 (27.1) | 2 083 (49.6) | 979 (23.3) |
| Workers | 1 343 (75.7) | 432 (24.3) | 49 (3.7) | 383 (28.6) | 672 (50.1) | 286 (21.3) |
| Retired | 736 (68.0) | 346 (32.0) | 10 (1.4) | 133 (18.2) | 356 (48.6) | 243 (33.2) |
| Office clerks | 509 (69.4) | 224 (30.6) | 26 (5.1) | 157 (31.0) | 242 (47.8) | 107 (21.1) |
| Students | 249 (76.4) | 77 (23.6) | 13 (5.2) | 76 (30.5) | 119 (47.8) | 54 (21.7) |
| Self-employed | 213 (71.2) | 86 (28.8) | 8 (3.8) | 61 (28.6) | 104 (48.8) | 48 (22.5) |
| Civil servants | 144 (66.4) | 73 (33.6) | 2 (1.4) | 37 (25.9) | 71 (49.7) | 35 (24.5) |
| Drivers | 113 (72.4) | 43 (27.6) | 3 (2.7) | 33 (29.2) | 56 (49.6) | 24 (21.2) |
| Others<sup>b</sup> | 2 749 (72.8) | 1 027 (27.2) | 62 (2.3) | 508 (18.6) | 1 372 (50.1) | 858 (31.3) |
| Missing | 650 (79.1) | 172 (20.9) | 10 (1.5) | 172 (26.6) | 351 (54.3) | 124 (19.2) |
| Etiological groups, n (%) |  |  |  |  |  |  |
| Low falls | 3 221 (72.3) | 1 231 (27.7) | 65 (2.0) | 641 (20.0) | 1 657 (51.7) | 909 (28.3) |
| High falls | 3 571 (81.2) | 827 (18.8) | 139 (3.9) | 1 095 (30.7) | 1762 (49.4) | 707 (19.8) |
| Traffic accidents | 2 489 (70.6) | 1 035 (29.4) | 65 (2.6) | 564 (22.7) | 1 221 (49.2) | 696 (28.1) |
| Struck by falling objects | 681 (77.3) | 200 (22.7) | 39 (5.7) | 194 (28.6) | 320 (47.1) | 165 (24.3) |
| Sport related injuries | 103 (62.0) | 63 (38.0) | 1 (1.0) | 20 (19.4) | 43 (41.7) | 40 (38.8) |
| Others<sup>c</sup> | 552 (62.4) | 332 (37.6) | 13 (2.4) | 92 (16.7) | 262 (47.5) | 197 (35.8) |
| Missing | 301 (67.0) | 148 (33.0) | 2 (0.7) | 92 (31.0) | 161 (54.2) | 44 (14.8) |
| Level of injury, n (%) |  |  |  |  |  |  |
| Cervical | 6 339 (70.9) | 2 605 (29.1) | 124 (2.0) | 1 203 (19.0) | 3 351 (53.0) | 1 769 (28.0) |
| Thoracic | 1 905 (80.3) | 468 (19.7) | 73 (3.9) | 547 (28.9) | 844 (44.5) | 505 (26.6) |
| Lumbosacral | 2 374 (80.3) | 583 (19.7) | 117 (4.9) | 874 (36.9) | 1 074 (45.4) | 419 (17.7) |
| Multi-site | 161 (88.0) | 22 (12.0) | 7 (4.4) | 48 (30.0) | 77 (48.1) | 35 (21.9) |
| Cauda equina | 27 (93.1) | 2 (6.9) | 0 (0.0) | 3 (11.1) | 18 (66.7) | 6 (22.2) |
| Missing | 112 (41.8) | 156 (58.2) | 3 (2.8) | 23 (21.1) | 62 (56.9) | 24 (22.0) |
| Severity, n (%) |  |  |  |  |  |  |
| Complete quadriplegia | 1 274 (70.8) | 526 (29.2) | 66 (5.2) | 395 (31.1) | 638 (50.2) | 239 (18.8) |
| Complete paraplegia | 2 343 (80.3) | 576 (19.7) | 130 (5.6) | 799 (34.2) | 1 023 (43.8) | 512 (21.9) |
| Incomplete quadriplegia | 4 590 (70.8) | 1 893 (29.2) | 46 (1.0) | 688 (15.0) | 2 486 (54.3) | 1 404 (30.7) |
| Incomplete paraplegia | 2 562 (78.2) | 713 (21.8) | 81 (3.2) | 795 (31.1) | 1 206 (47.2) | 553 (21.7) |
| Missing | 149 (53.8) | 128 (46.2) | 1 (0.7) | 21 (14.6) | 73 (50.7) | 50 (34.7) |
| AIS grade<sup>d</sup>, n (%) |  |  |  |  |  |  |
| A | 2 824 (76.9) | 847 (23.1) | 157 (5.6) | 1 009 (35.8) | 1 259 (44.7) | 549 (19.5) |
| B | 825 (81.8) | 184 (18.2) | 48 (5.8) | 246 (30.0) | 421 (51.3) | 154 (18.8) |
| C | 1 937 (77.1) | 575 (22.9) | 52 (2.7) | 397 (20.6) | 1 057 (54.8) | 476 (24.7) |
| D | 5 117 (71.4) | 2 051 (28.6) | 62 (1.2) | 1 012 (19.8) | 2 571 (50.4) | 1 521 (29.8) |
| E | 162 (65.3) | 86 (34.7) | 5 (3.1) | 26 (16.1) | 96 (59.6) | 39 (24.2) |
| Missing | 53 (36.3) | 93 (63.7) | 0 (0.0) | 8 (16.3) | 22 (44.9) | 19 (38.8) |

<sup>a</sup> "Surgery timing" represented the time from injury to surgery (h=hours, d=days). <sup>b</sup> "Others" included "Teachers", "Unemployed" and "Freelances". <sup>c</sup> "Others" included "Injuries caused by others", "Work related injuries", "Sharp injuries", "Massage related injuries", "Electric shock injuries", "Gunshot wounds", "Crush injuries", and "Iatrogenic injuries". <sup>d</sup> AIS grade A represents complete impairment with no motor or sensory function below the level of injury; grades B–D represent progressively less severe impairment; AIS grade E represents normal motor and sensory examinations, but still may have abnormal reflexes or other neurologic phenomena. TSCI=traumatic spinal cord injury. AIS=American Spinal Injury Association (ASIA) Impairment Scale.

### Glucocorticoids

2 084 (14.1% of 14 754) patients were treated with MPSS/MP at a high dose (≥500 mg), of which 641 patients (30.8% of 2 084) received it within 8 hours, followed by 1 443 (69.2% of 2 084) at least 8 hours after TSCI. There were more than one third (5 352, 36.3% of 14 754) patients receiving a normal dose of MPSS/MP (<500 mg), of which most patients (5 012, 93.6% of 5 352) received it continuously and few (340, 6.4% of 5 352) intermittently (Figure 2 and Table supplement 3).

### Neurotrophic Drugs, Dehydrants and Cathartics

About two thirds (9 590, 65.0% of 14 754) of the patients were treated with neurotrophic drugs, and the top three categories were GM-1 ganglioside (4 979, 51.9%), mouse nerve growth factor (2 785, 29.0%) and mecobalamin (2 580, 26.9%). Among cases with dehydrants (8 898, 60.3% of 14 754) , mannitol (7 558, 84.9%) was the most commonly used. Glycerin or glycerine enema (1 434, 95.1%) accounted for the highest proportion of cases with cathartics (1 508, 10.2% of 14 754). (Table supplement 4)

### Economic Burden and length of hospital stay

The mean total costs among the 10 945 acute TSCI cases were CNY ¥ 71.0k (USD $ 11.5k), with daily costs of ¥ 4.4k ($ 0.7k). Both total and daily costs decreased gradually with age. Total costs were highest in students (¥ 79.6k, $ 12.9k) but daily costs were highest in workers (¥ 4.4k, $ 0.7k) considering occupations categories. Total costs were highest in patients struck by falling objects (¥ 95.9k, $ 15.5k) while daily costs was highest in high falls (¥ 4.9k, $ 0.8k) considering etiological groups. Total and daily costs were highest in cauda equina (¥ 94.1k, $ 15.2k; ¥ 5.1k, $ 0.8k) considering injury level. Both total and daily costs increased with the ncreasing of AIS grade (E to A). The average length of hospital stay was 19.9 (SD= 26.2) days. (Tables 3 and Table supplement 5)

**Table 3:** The costs among acute TSCI patients during hospitalization.

| Category | Number of<br>patients, n | Total costs | Daily costs |
| --- | --- | --- | --- |
|  |  | (CNY/USD, thousand) | (CNY/USD, thousand) |
|  |  | Mean (SD) | Mean (SD) |
| Gender |  |  |  |
| Men | 8 304 | ¥ 73.6 (72.3), \$ 11.9 (11.7) | ¥ 4.4 (3.4), \$ 0.7 (0.6) |
| Women | 2 641 | ¥ 62.6 (65.1), \$ 10.1 (10.5) | ¥ 4.2 (3.5), \$ 0.7 (0.6) |
| Total | 10 945 | ¥ 71.0 (70.8), \$ 11.5 (11.4) | ¥ 4.4 (3.4), \$ 0.7 (0.6) |
| Age groups |  |  |  |
| 15 years~ | 602 | ¥ 83.5 (100.9), \$ 13.5 (16.3) | ¥ 4.6 (3.5), \$ 0.7 (0.6) |
| 25 years~ | 1 223 | ¥ 78.5 (70.9), \$ 12.7 (11.4) | ¥ 4.5 (3.5), \$ 0.7 (0.6) |
| 35 years~ | 1 928 | ¥ 75.6 (79.7), \$ 12.2 (12.9) | ¥ 4.5 (3.4), \$ 0.7 (0.5) |
| 45 years~ | 3 073 | ¥ 73.1 (65.2), \$ 11.8 (10.5) | ¥ 4.4 (3.5), \$ 0.7 (0.6) |
| 55 years~ | 2 439 | ¥ 65.9 (58.5), \$ 10.6 (9.4) | ¥ 4.2 (3.5), \$ 0.7 (0.6) |
| ≥ 65 years | 1 680 | ¥ 59.0 (70.7), \$ 9.5 (11.4) | ¥ 4.1 (3.2), \$ 0.7 (0.6) |
| Occupations |  |  |  |
| Farmers | 4 215 | ¥ 64.7 (54.6), \$ 10.4 (8.8) | ¥ 4.3 (3.3), \$ 0.7 (0.5) |
| Workers | 1 387 | ¥ 79.0 (75.7), \$ 12.7 (12.2) | ¥ 4.4 (4.0), \$ 0.7 (0.6) |
| Retired | 769 | ¥ 55.5 (52.5), \$ 9.0 (8.5) | ¥ 4.0 (3.1), \$ 0.6 (0.5) |
| Office clerks | 541 | ¥ 70.6 (72.7), \$ 11.4 (11.7) | ¥ 4.4 (4.1), \$ 0.7 (0.7) |
| Students | 282 | ¥ 79.6 (124.4), \$ 12.9 (20.1) | ¥ 4.2 (3.8), \$ 0.7 (0.6) |
| Self-employed | 229 | ¥ 61.3 (52.3), \$ 10.7 (9.1) | ¥ 3.8 (2.9), \$ 0.6 (0.5) |
| Civil servants | 179 | ¥ 63.1 (60.2), \$ 9.9 (8.4) | ¥ 3.9 (3.9), \$ 0.6 (0.6) |
| Drivers | 134 | ¥ 74.5 (97.0), \$ 12.0 (15.7) | ¥ 3.8 (2.7), \$ 0.6 (0.4) |
| Others<sup>a</sup> | 2 759 | ¥ 79.0 (85.3), \$ 12.8 (13.8) | ¥ 4.5 (3.2), \$ 0.7 (0.5) |
| Missing | 450 | ¥ 83.9 (60.7), \$ 13.5 (9.8) | ¥ 5.6 (3.4), \$ 0.9 (0.5) |
| Etiological groups |  |  |  |
| Low falls | 3 260 | ¥ 54.7 (49.2), \$ 8.8 (8.0) | ¥ 3.9 (3.3), \$ 0.6 (0.5) |
| High falls | 3 459 | ¥ 81.2 (73.6), \$ 13.1 (11.9) | ¥ 4.9 (3.5), \$ 0.8 (0.6) |
| Traffic accidents | 2 705 | ¥ 73.8 (74.5), \$ 11.9 (12.0) | ¥ 4.3 (3.3), \$ 0.7 (0.5) |
| Struck by falling objects | 657 | ¥ 95.9 (110.8), \$ 15.5 (17.9) | ¥ 4.8 (4.0), \$ 0.8 (0.6) |
| Sport related injuries | 117 | ¥ 42.4 (42.0), \$ 6.8 (6.8) | ¥ 4.0 (4.2), \$ 0.7 (0.7) |
| Others<sup>b</sup> | 573 | ¥ 64.4 (68.5), \$ 10.4 (11.1) | ¥ 3.6 (2.9), \$ 0.6 (0.5) |
| Missing | 174 | ¥ 74.2 (51.3), \$ 12.0 (8.3) | ¥ 5.3 (3.3), \$ 0.9 (0.5) |
| Level of injury |  |  |  |
| Cervical | 6 594 | ¥ 65.9 (68.4), \$ 10.6 (11.1) | ¥ 4.2 (3.3), \$ 0.7 (0.5) |
| Thoracic | 1 783 | ¥ 94.1 (90.5), \$ 15.2 (14.6) | ¥ 5.0 (3.8), \$ 0.8 (0.6) |
| Lumbosacral | 2 242 | ¥ 68.2 (56.9), \$ 11.0 (9.2) | ¥ 4.4 (3.7), \$ 0.7 (0.6) |
| Multi-site | 132 | ¥ 72.4 (43.2), \$ 11.7 (7.0) | ¥ 5.0 (2.7), \$ 0.8 (0.4) |
| Cauda equina | 37 | ¥ 94.1 (39.6), \$ 15.2 (6.4) | ¥ 5.1 (1.7), \$ 0.8 (0.3) |
| Missing | 157 | ¥ 52.5 (59.1), \$ 8.5 (9.5) | ¥ 3.5 (2.7), \$ 0.6 (0.4) |
| Severity |  |  |  |
| Complete quadriplegia | 1 417 | ¥ 92.5 (102.0), \$ 14.9 (16.5) | ¥ 4.8 (3.3), \$ 0.8 (0.5) |
| Complete paraplegia | 2 248 | ¥ 93.0 (89.2), \$ 15.0 (14.4) | ¥ 5.1 (4.2), \$ 0.8 (0.7) |
| Incomplete quadriplegia | 4 702 | ¥ 58.6 (50.7), \$ 9.5 (8.2) | ¥ 4.0 (3.2), \$ 0.6 (0.5) |
| Incomplete paraplegia | 2 467 | ¥ 62.7 (53.9), \$ 10.1 (8.7) | ¥ 4.2 (3.0), \$ 0.7 (0.5) |
| Missing | 111 | ¥ 57.1 (51.2), \$ 9.2 (8.3) | ¥ 3.7 (2.9), \$ 0.6 (0.5) |
| AIS grade<sup>d</sup> |  |  |  |
| A | 2 958 | ¥ 96.4 (101.5), \$ 15.6 (16.4) | ¥ 5.1 (4.0), \$ 0.8 (0.6) |
| B | 731 | ¥ 92.5 (71.7), \$ 14.9 (11.6) | ¥ 5.1 (3.8), \$ 0.8 (0.6) |
| C | 1 831 | ¥ 75.9 (56.2), \$ 12.3 (9.1) | ¥ 4.5 (3.0), \$ 0.7 (0.5) |
| D | 5 208 | ¥ 52.4 (43.5), \$ 8.5 (7.0) | ¥ 3.8 (3.1), \$ 0.6 (0.5) |
| E | 171 | ¥ 49.5 (53.6), \$ 8.0 (8.7) | ¥ 3.3 (3.0), \$ 0.5 (0.5) |
| Missing | 46 | ¥ 70.4 (76.7), \$ 11.4 (12.4) | ¥ 4.1 (2.3), \$ 0.7 (0.4) |

<sup>a</sup> "Others" included "Teachers", "Unemployed" and "Freelances". <sup>b</sup> "Others" included "Injuries caused by others", "Work related injuries", "Sharp injuries", "Massage related injuries", "Electric shock injuries", "Gunshot wounds", "Crush injuries", and "Iatrogenic injuries". <sup>c</sup> AIS grade A represents complete impairment with no motor or sensory function below the level of injury; grades B–D represent progressively less severe impairment; AIS grade E represents normal motor and sensory examinations, but still may have abnormal reflexes or other neurologic phenomena. TSCI=traumatic spinal cord injury. AIS=American Spinal Injury Association (ASIA) Impairment Scale. CNY=Chinese Yuan. USD=USA dollar. SD=standard deviation.

Between 2013 and 2018, the total costs for acute TSCI decreased (APC= -4.8%, 95% CI: -6.2 o -3.4). The daily costs did not change significantly (APC= 0.5%, 95% CI: -1.2 to 2.2), the same for both men (APC= 0.3%, 95% CI: -1.8 to 2.4) and women (APC= 1.7%, 95% CI: -1.6 o 5.2). The daily costs of each age group did not change significantly, except for patients aged ≥65 years with a increase (APC= 2.9%, 95% CI: 0.1 to 5.9). In addition, the average ength of hospital stay decreased (APC= -4.5, 95% CI: -8.1 to -0.6). (Table supplement 5 and Table supplement 6)

## Discussion

To our knowledge, this is the first national-level study of the epidemiological and clinical features, treatment status and economic burden of TSCI in China. The study was a retrospective multicentre hospital-based study of TSCI from 2013 to 2018. We found that the percentage of TSCI in hospitalized patients and the percentage of TSCI in hospitalized patients through the orthopaedic departments did not change significantly overall, but the percentage of TSCI in both hospitalized patients and hospitalized patients through the orthopaedic departments increased when the number of TSCI per year was greater than 140. We described the treatment status with regard to: surgery, MPSS/MP, neurotrophic drugs, dehydrants and cathartics in detail. The total costs for acute TSCI decreased, while the daily costs did not change significantly.

The epidemiological features, such as gender (male: 75.3%) and average age (50.0 years) were consistent with other previous area studies of TSCI in China *(Yuan et al., 2018)*. Our results showed that TSCI patients were more likely to be farmers (37.7%), which was different from the epidemiological studies in Turkey and Mexico *(Güzelküçük et al., 2015; Zá rate-Kalfópulos et al., 2016)*. The higher number of farmers suffering TSCI in China compared with other countries might reflect the type of work farmers in China are undertaking, including high-risk construction, which increases the risk of falls from elevated heights, which made up 29.8% of all TSCI *(National Bureau of Statistics, 2021)*.

From 2013 to 2018, the percentage of TSCI in hospitalized patients and the percentage of TSCI in hospitalized patients through the orthopaedic departments did not change significantly overall. This finding was similar with the trend of age-standardized incidence rates of spinal cord injury in China reported in the GBD study *(GBD 2016 Traumatic Brain Injury and Spinal Cord Injury Collaborators, 2019)*. The percentage of TSCI in both hospitalized patients and hospitalized patients through the orthopaedic departments increased when the number of TSCI per year was greater than 140, which might reflect a changing trend n China of TSCI patients being treated in hospitals with more TSCI expertise. Transferring patients who suffer a TSCI into hospitals with speciality in TSCI is likely to improve the quality of the patients care, and might also help reduce the burden on the large non-specialist hospitals. In China, the over-burdening of the large non-specialist hospitals has prevented optimal care from being provided for TSCI, such as early treatment *(National Health Commission of the People’s Republic of China, 2019)*.

One of the recommended early treatment for TSCI is surgery *(Robert R Hansebout, 2021)*, and a recent pooled analysis including 1 548 patients showed that surgery decompression within 24 hours of acute spinal cord injury was associated with improved sensorimotor recovery *(Badhiwala et al., 2021)*. In our study, about 3.0% of 10 882 patients underwent surgery received surgery less than 24 hours after injury. Irrespective of the preference of clinicians on the use of early surgery, most hospitals do not have capacity to complete early surgery of TSCI because they lacked experienced surgeons, imaging and laboratory equipment, and preanesthetic assessments *(Thompson et al., 2018)*.

Another key area of managing TSCI is pharmacological interventions. Current pharmacological treatment principally relies on the off-label use of MPSS/MP, which remains contentious *(Robert R Hansebout, 2021)*. Based upon the available evidence, the American Association of Neurological Surgeons and the Congress of Neurological Surgeons (AANS/CNS) stated that "administration of methylprednisolone (MP) for the treatment of acute spinal cord injury is not recommended" in 2013, and concluded that "there is nsufficient evidence to make a recommendation" of using MP for treatment of patients with horacic and lumbar fractures and spinal cord injury in 2019 *(Hurlbert et al., 2013; Arnold et al., 2019)*. AOSpine organization on the other hand suggested a high-dose of MPSS/MP within 8 hours of acute SCI as a treatment option in their 2017 guideline *(Fehlings et al., 2017)*. A retrospective population-based cohort study showed that 61.5% (rang from 40.7 to 75.7) patients in South Korea between 2007 and 2017 received high-dose MPSS/MP within 8 hours of acute spinal cord injury *(Choi et al., 2020)*. In our study, we used 500mg of MPSS/MP as a threshold, based on the user guide, and found that 2 084 (14.1% of 14 754) TSCI patients were treated with a MPSS/MP of ≥500 mg, with only 641 (4.3% of 14 754) patients receiving it within 8 hours. The low rates of MPSS/MP use might reflect the conflicting recommendations in guidelines, the ongoing controversy in clinical management, professionals treating TSCI patients *(Robert R Hansebout, 2021; Arnold et al., 2019)*. Future high-quality randomized controlled trials or comprehensive observational studies based on arge-scale electronic medical records are therefore needed, to explore the benefit of glucocorticoids treatment and help inform clinical care *(Liu et al., 2019)*.

In addition to the low rates of early surgery and glucocorticoids use identified in this study, here was also found to be common use of treatments not part of recommended usual care in reating TSCI, such as neurotrophic drugs, dehydrants, and cathartics. Moreover, the AANS/CNS stated that "administration of GM-1 ganglioside (Sygen) for the treatment of acute SCI was not recommended" in 2013 and "there is insufficient evidence to make a recommendation" to use GM-1 ganglioside for patients with thoracic and lumbar fractures and spinal cord injury in 2019 *(Hurlbert et al., 2013; Arnold et al., 2019)*. In this study, 9 590 (65.0%) patients used neurotrophic drugs, of which 4 979 (51.9%) patients used GM-1 ganglioside. The high rates of neurotrophic drugs in China might reflect the insufficient high-quality evidence to recommend for or against it.

Considering the costs related to acute TSCI, we found that the total costs on average were CNY ¥ 71.0k (USD $ 11.5k), which was 3.9 times that of disposable income per capita of 2013 (¥ 18.3k) (from the National Bureau of Statistics of the People’s Republic of China) in China *(National Bureau of Statistics, 2021)*. These costs are not only a large burden for spinal cord injuries (3 739 610) and the second highest number of new cases (98 226) in 2016 worldwide based on the 2016 GBD study *(GBD 2016 Traumatic Brain Injury and Spinal Cord Injury Collaborators, 2019)*. The total costs decreased but the daily costs did not change significantly during the same period, which was consistent with the reduction in the average ength of hospital stay. On the one hand, the cause of hospital stay reduction might be due to most hospitals improving the diagnosis and treatment or/and simplifying the process in and out of the hospital in China *(National Health Commission of the People’s Republic of China, 2019)*. On the other hand, it might be a reflection of patients leaving the hospital earlier to save money, rather than due to improved care leading to early discharge. The early discharge from hospital might also lead to deep wound infection, wound dehiscence, and deep venous hrombosis *(Otero et al., 2016)*.

Although this study was the largest known study of TSCI cases in China, it still possessed some limitations. First, it was not a population-based design, which meant that we could not calculate the incidence and prevalence rates of TSCI from the whole population. Another imitation in this study was the representative of sample. Although many hospitals were chosen across China, we did not use strict sampling rule so that our study is not a national representative one. Third, we only described the use of MPSS/MP in the part of glucocorticoids usage, without hydrocortisone and dexamethasone, due to the uncertainty whether other glucocorticoids were used in the treatment of TSCI itself.

## Conclusion

This study revealed the epidemiological and clinical features of TSCI that occurred in China from 2013 to 2018. We found that the percentage of TSCI in hospitalized patients and the percentage of TSCI in hospitalized patients through the orthopaedic departments did not change significantly, but both these percentages increased when the number of TSCI per year was greater than 140. We described the treatment status about surgery, MPSS/MP, neurotrophic drugs, dehydrants and cathartics. The total costs for TSCI decreased, but the daily costs did not change significantly. Further studies should focus on assessing the efficacy and safety of current treatment strategies.

## Data Availability

The data are available from the corresponding author on reasonable request.

## Acknowledgments

We thank Ping Yu from Library of Tianjin Medical University for developing the search strategies for TSCI. We thank Jiaxiao Shi, Xuanhao Fu, Wenxiang Li, Shibo Zhu, Linlin Shi, Weixiao Liu, Shenghui Shang, Shiyang Yuan, Guowang Li, Chao Sun, Huiquan Duan, Siyue Jia, Chaoyu Wang, Ziqian Xiang, Chao Li, Yigang Lv and Hao Yu for data collection. We hank Wei Li from Zhengzhou Orthopaedics Hospital, Yi Zhang from The First Affiliated Hospital of Zhengzhou University, Dantong Sun from The Eighth People’s Hospital of Shenyang, Zanjing Zhai from Shanghai Ninth People’s Hospital, Shanghai Jiao Tong University School of Medicine, Bo Wu from Shenyang Orthopaedics Hospital, Xueming Chen from Beijing Luhe Hospital, Affiliated to Capital Medical University, Xianze Sun from The Third Hospital of Shijiazhuang, Zaixian Long from Xiushan People’s Hospital, Si Yin from The First Affiliated Hospital of Xi’An Jiaotong University, Shenglin Zhang from Jinhu County People’s Hospital. Zhihua Wang from People’s Hospital of Wuzhi County. Wei Ma and Xing Dai from Hanyin county people’s hospital, Jidong Li from Hospital of Jingxing County, Dazhi Yang from Shenzhen People’s Hospital, The Second Clinical Medical College of Jinan University, Jiancheng Zeng from West China Hospital, Sichuan University, and Deguo Teng from The Second People’s Hospital of Shifang for data management.

## Contributors

SQF and HXZ designed the study and obtained funding. HXZ, YFL, LXC, YK and JHW did data analysis and the preparation of the manuscript. LL, ZWC and CZ did figures design. All authors read and approved the final version.

## Data sharing

The data are available from the corresponding author on reasonable request.

## Supplementary Information

**Figure supplement 1.**
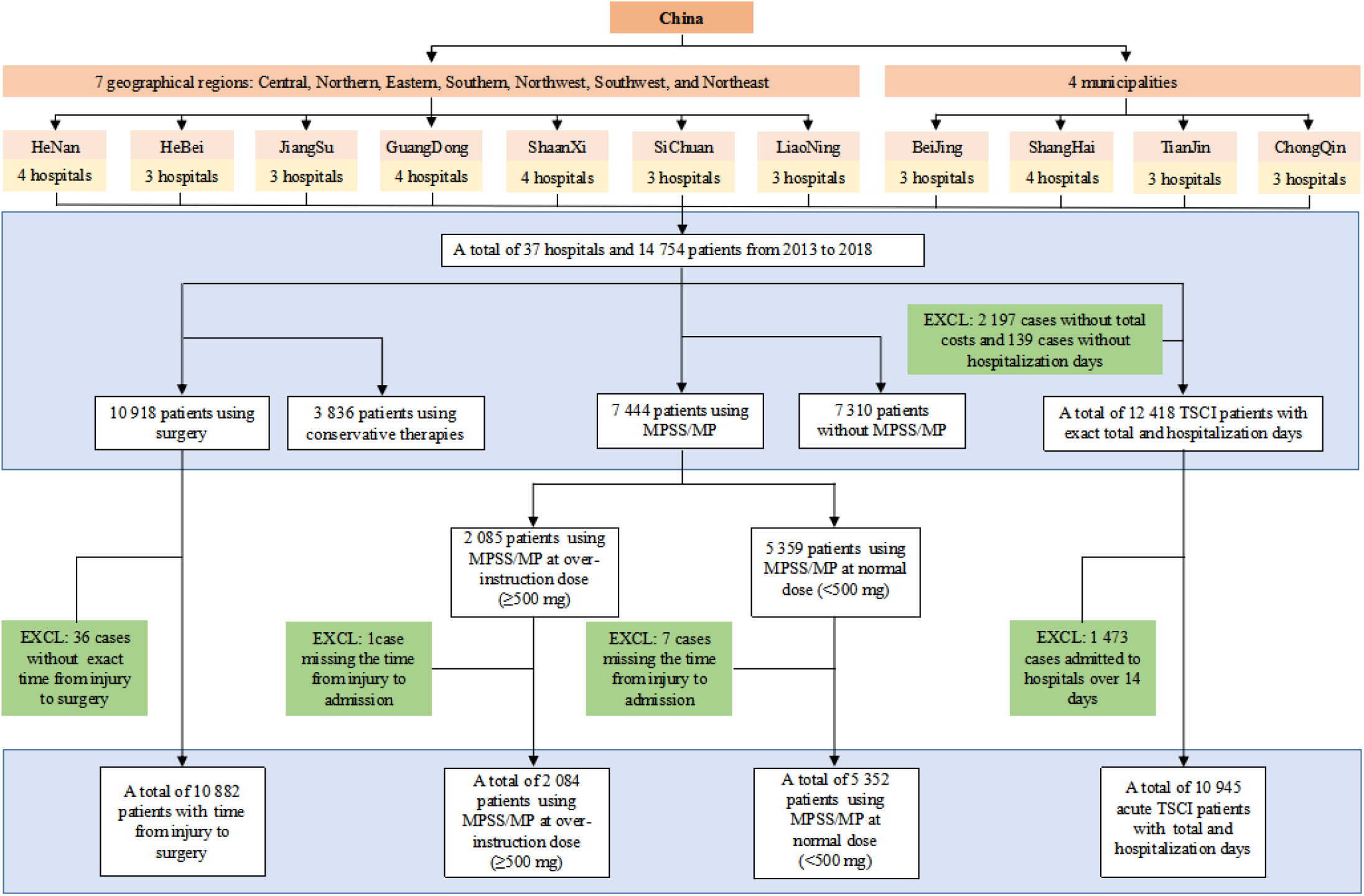
Flow chart of case identification. In the end, a total of 14 754 patients from 37 hospitals in China were included in this study from 2013 to 2018. And the final number of cases included in the "surgery timing", the use of MPSS/MP, and the costs of acute TSCI during hospitalization were shown in the above figure. TSCI=traumatic spinal cord injury . EXCL=excluded. MPSS/MP=methylprednisolone sodium succinate/methylprednisolone.

**Figure supplement 2.**
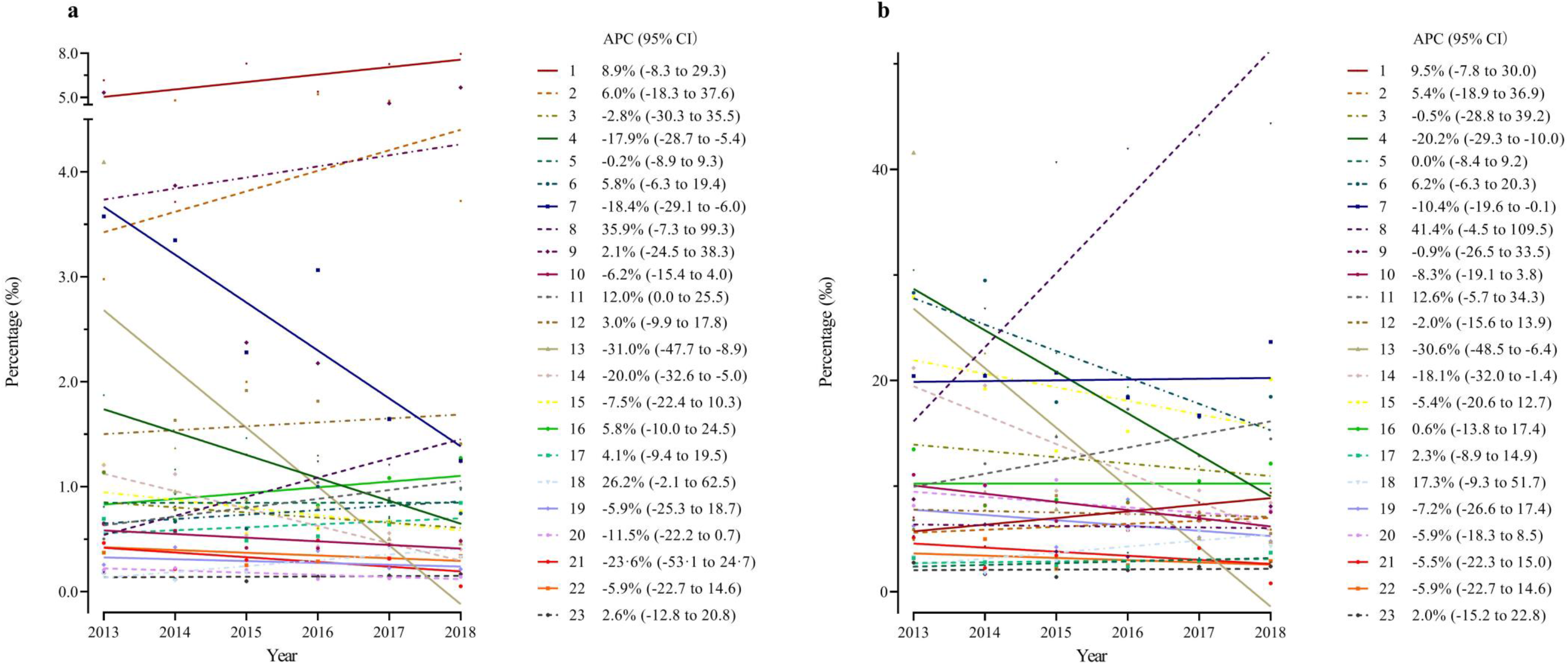
Trends in the percentage among TSCI patients of 23 hospitals from 2013 to 2018. (a) Trends in the percentage of TSCI in hospitalized patients of each hospital from 23 hospitals. (b) Trends in the percentage of TSCI in hospitalized patients through the orthopaedic departments of each hospital from 23 hospitals. TSCI=traumatic spinal cord injury. APC=annual percentage change. 95% CI=95% confidence interval.

**Figure supplement 3.**
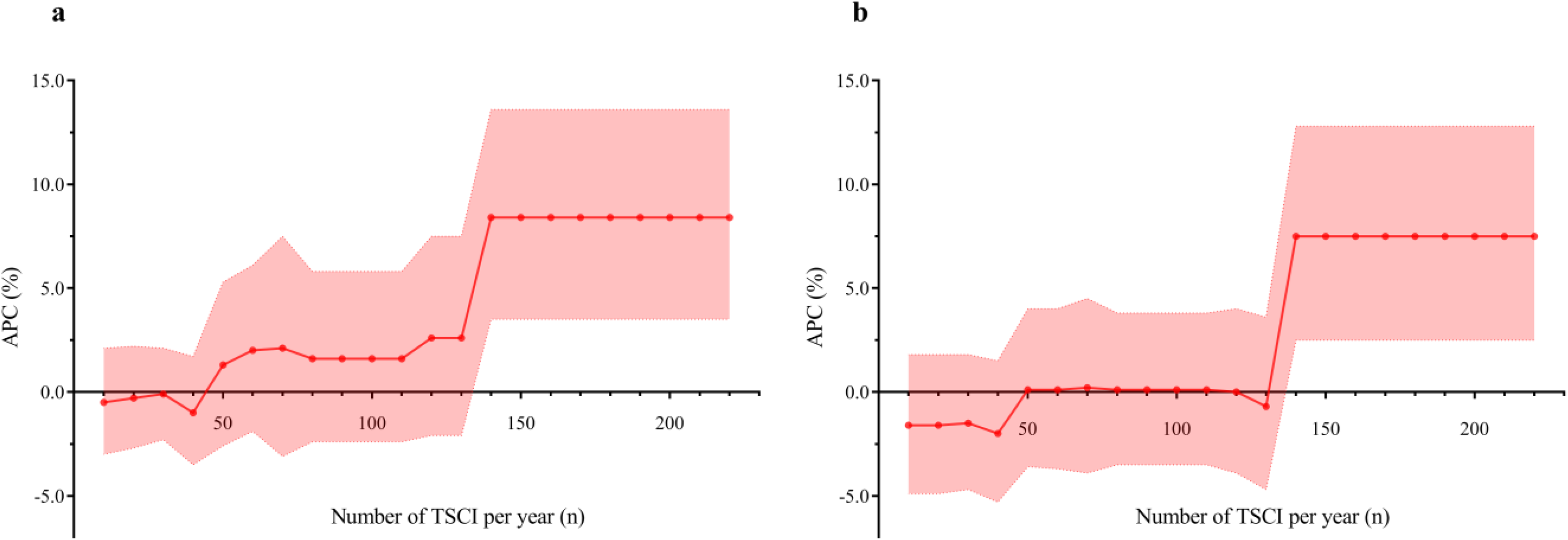
The trends of APC in the percentage among TSCI patients from 23 hospitals. A The trend of APC in the percentage among TSCI patients in hospitalized patients of different number of TSCI per year. (b) The trend of APC in the percentage among TSCI patients in hospitalized patients through the orthopaedic departments of different number of TSCI per year. The value of the horizontal axis represented the annual TSCI patients admitted. The APC corresponding to the points referred to the comprehensive calculation of APC from 2013 to 2018 of all eligible hospitals whose annual TSCI patients were more than the corresponding number of horizontal axis in 23 hospitals. Shaded regions represent 95% confidence interval. APC=annual percentage change. TSCI=traumatic spinal cord injury.

**Table supplement 1.** American Spinal Injury Association (ASIA) Impairment Scale (AIS) grade.

| AIS grade | Category | Description |
| --- | --- | --- |
| A | Complete | No motor or sensory function is preserved in the sacral segments S4–S5 |
| B | Incomplete | Sensory function preserved but not motor function is preserved below the neurological level and includes the sacral segments S4–S5 |
| C | Incomplete | Motor function is preserved below the neurological level, and more than half of key muscles below the neurological level have a muscle grade less than 3 |
| D | Incomplete | Motor function is preserved below the neurological level, and at least half of key muscles below the neurological level have a muscle grade of 3 or more |
| E | Normal | Motor and sensory function are normal, but still may have abnormal reflexes or other neurologic phenomena<sup>a</sup>. |

**Table supplement 2.** Trends in the percentage of different subgroups among TSCI patients from 2013 to 2018.

| Category | 2013 | 2014 | 2015 | 2016 | 2017 | 2018 | APC (95% CI) |
| --- | --- | --- | --- | --- | --- | --- | --- |
| **TSCI patients/Hospitalized patients** |  |  |  |  |  |  |  |
| Hospital type |  |  |  |  |  |  |  |
| General hospital | 0.000677437 | 0.000731737 | 0.000650481 | 0.000650376 | 0.000584477 | 0.000574924 | -4.2 (-7.2, -1.0) |
| Orthopaedic hospital | 0.004146887 | 0.004100949 | 0.003582044 | 0.004432452 | 0.004789523 | 0.004711542 | 3.8 (-2.2, 10.3) |
| Economic level |  |  |  |  |  |  |  |
| Above the PCDI | 0.000764280 | 0.000854571 | 0.000837578 | 0.000813096 | 0.000837774 | 0.000864237 | 1.5 (-1.1, 4.2) |
| Below the PCDI | 0.001961198 | 0.001901074 | 0.001352715 | 0.002064160 | 0.001701302 | 0.001434080 | -4.1 (-14.5, 7.6) |
| **TSCI patients/Orthopaedic patients** |  |  |  |  |  |  |  |
| Hospital type |  |  |  |  |  |  |  |
| General hospital | 0.009496961 | 0.010165120 | 0.008524518 | 0.008521055 | 0.007753479 | 0.007722081 | -5.1 (-8.4, -1.7) |
| Orthopaedic hospital | 0.006207775 | 0.006087330 | 0.005567732 | 0.006757954 | 0.007892531 | 0.007828871 | 6.3 (-0.1, 13.1) |
| Economic level |  |  |  |  |  |  |  |
| Above the PCDI | 0.007594290 | 0.008133067 | 0.007529879 | 0.007149736 | 0.007625364 | 0.007885594 | -0.2 (-3.3, 3.1) |
| Below the PCDI | 0.010551825 | 0.010410185 | 0.006861413 | 0.010631962 | 0.008536560 | 0.007279922 | -5.6 (-16.5, 6.8) |

**Table supplement 3.** The use of MPSS/MP among TSCI patients during hospitalization.

| Category<sup>a</sup> | accumulated time≤2 d | 3 d ≤accumulated time≤5 d | accumulated time≥6 d | No accumulated time<sup>b</sup> | Total |
| --- | --- | --- | --- | --- | --- |
| Use of MPSS/MP at over-instruction dose (≥500 mg) |  |  |  |  |  |
| ≤8 h | -- | -- | -- | 641 | 641 |
| >8 h | -- | -- | -- | 1 443 | 1 443 |
| Total | -- | -- | -- | 2 084 | 2 084 |
| Use of MPSS/MP with normal instruction dose (<500 mg) |  |  |  |  |  |
| Total |  |  |  |  | 5 352 |
| Continuous use of MPSS/MP with normal instruction dose |  |  |  |  |  |
| ≤1.6 d | 495 | 439 | 320 | -- | 1 254 |
| 1.6-9.0 d | 897 | 923 | 629 | -- | 2 449 |
| ≥9.0 d | 505 | 553 | 251 | -- | 1 309 |
| Total | 1 897 | 1 915 | 1 200 | -- | 5 012 |
| Intermittent use of MPSS/MP with normal instruction dose |  |  |  |  |  |
| ≤1.6 d | 15 | 63 | 75 | -- | 153 |
| 1.6-9.0 d | 11 | 62 | 86 | -- | 159 |
| ≥9.0 d | 4 | 9 | 15 | -- | 28 |
| Total | 30 | 134 | 176 | -- | 340 |

<sup>a</sup> The rows represented the start time of using MPSS/MP (h=hours, d=days), the columns represented the accumulated time of medication (d=days), and the figures represented the number of TSCI patients. <sup>b</sup> There is no accumulated time about use of MPSS/MP at over-instruction dose (≥500 mg). TSCI=traumatic spinal cord injury. MPSS/MP=methylprednisolone sodium succinate/methylprednisolone.

**Table supplement 4.**
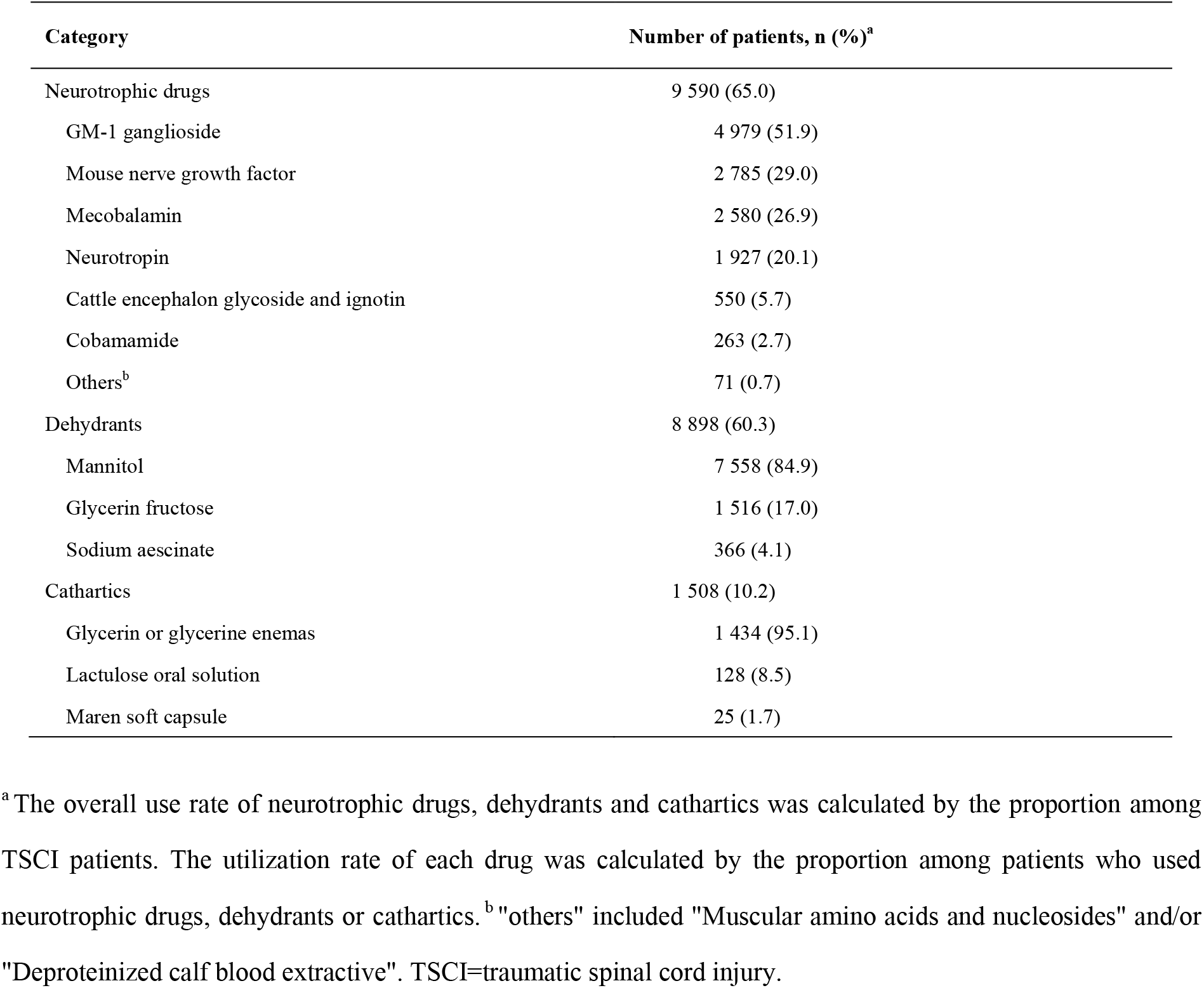
The use of neurotrophic drugs, dehydrants and cathartics among TSCI patients during hospitalization.

**Table supplement 5.** Trends in the length of hospital stay among acute TSCI patients from 2013 to 2018a.

| Category | Overall | 2013 | 2014 | 2015 | 2016 | 2017 | 2018 | APC (95% CI) |
| --- | --- | --- | --- | --- | --- | --- | --- | --- |
| Gender, Mean (SD) |  |  |  |  |  |  |  |  |
| Men | 20.4 (27.1) | 21.9 (20.7) | 23.1 (40.1) | 20.1 (23.7) | 21.2 (25.8) | 19.6 (28.6) | 17.7 (17.9) | -4.2 (-7.5, -0.8) |
| Women | 18.5 (23.2) | 19.4 (16.8) | 20.1 (19.5) | 20.4 (26.4) | 19.9 (25.4) | 18.1 (30.3) | 14.6 (15.0) | -4.9 (-10.7, 1.3) |
| Total | 19.9 (26.2) | 21.3 (19.8) | 22.4 (36.5) | 20.1 (24.3) | 20.9 (25.7) | 19.3 (29.0) | 16.8 (17.2) | -4.5 (-8.1, -0.6) |
| Age groups, Mean (SD) |  |  |  |  |  |  |  |  |
| 15 years~ | 22.4 (31.6) | 20.9 (14.9) | 25.0 (26.2) | 20.5 (24.0) | 23.2 (22.8) | 26.5 (64.6) | 19.0 (16.2) | -0.5 (-9.5, 9.3) |
| 25 years~ | 21.5 (25.3) | 25.8 (29.4) | 25.2 (29.8) | 21.9 (30.2) | 20.2 (19.1) | 19.6 (22.2) | 18.8 (21.6) | -6.7 (-8.8, -4.5) |
| 35 years~ | 21.1 (35.1) | 21.8 (22.2) | 24.0 (62.8) | 21.6 (28.7) | 21.0 (27.6) | 18.7 (21.8) | 19.4 (23.7) | -3.8 (-7.4, -0.1) |
| 45 years~ | 20.9 (25.9) | 20.6 (14.7) | 22.5 (23.9) | 22.1 (27.4) | 21.8 (29.0) | 20.8 (32.4) | 18.0 (18.5) | -2.6 (-7.1, 2.2) |
| 55 years~ | 18.6 (19.9) | 20.7 (19.2) | 21.7 (27.6) | 16.8 (14.5) | 20.6 (22.5) | 16.7 (20.0) | 16.2 (12.5) | -5.0 (-11.1, 1.5) |
| ≥ 65 years | 16.9 (20.3) | 19.3 (19.1) | 16.9 (14.6) | 18.1 (17.6) | 19.5 (26.6) | 18.2 (27.1) | 12.5 (9.5) | -5.2 (-14.0, 4.5) |

**Table supplement 6.**
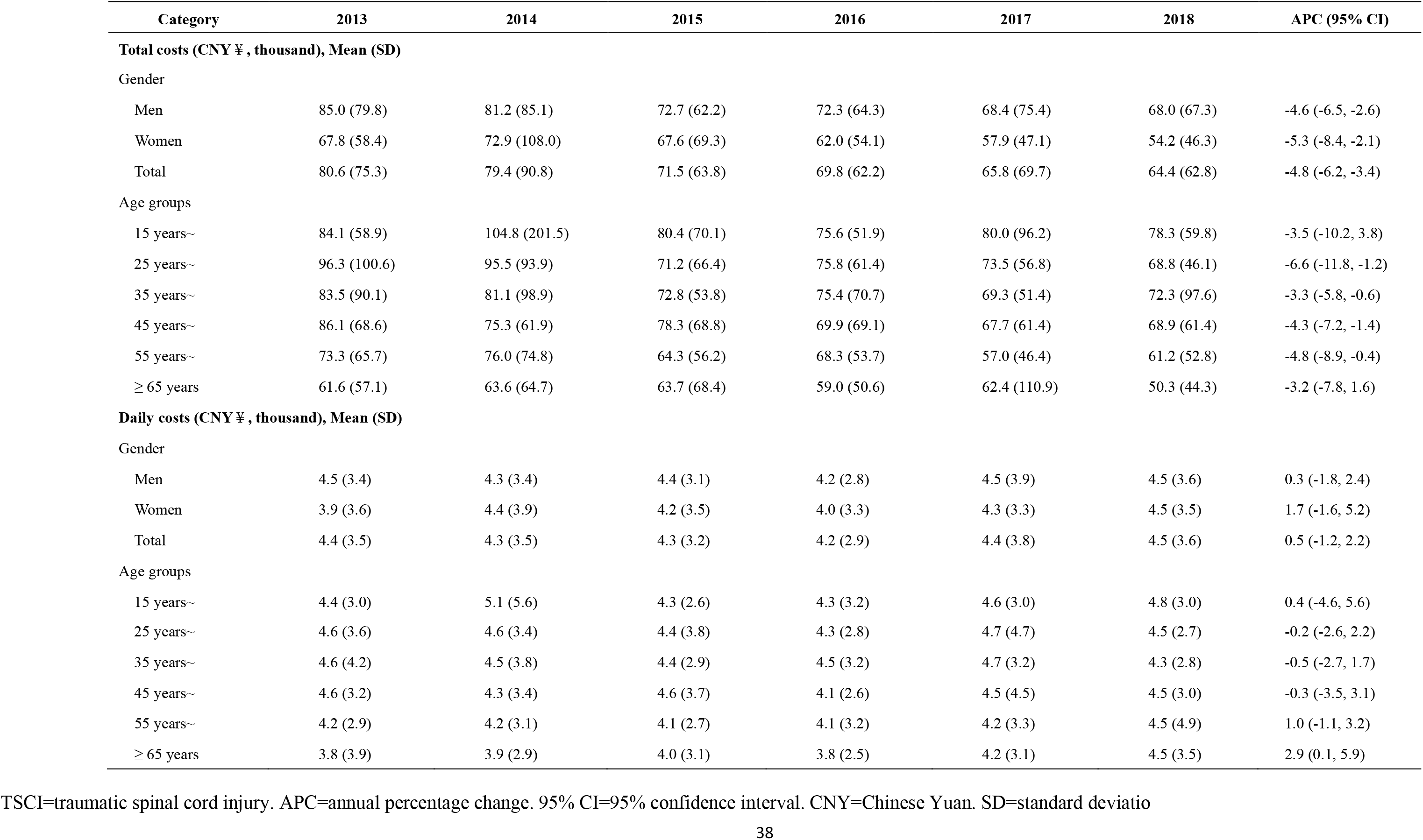
Trends in the costs among acute TSCI patients from 2013 to 2018.

